# EMT, angiogenesis, and cell-cycle pathways are significantly impacted in oral cancer patients, among whom a large fraction may potentially respond to immune checkpoint therapy

**DOI:** 10.1101/2024.11.09.24317044

**Authors:** Debodipta Das, Arindam Maitra, Chinmay K. Panda, Sandip Ghose, Bidyut Roy, Rajiv Sarin, Partha P. Majumder

## Abstract

**Background:** Oral squamous cell carcinoma of the gingivo-buccal region (OSCC-GB) has the highest incidence among males and the second highest overall among all cancers in India, emphasizing the need for precise molecular classifications to guide personalized therapy.

**Methods:** We performed bulk RNA sequencing on tumor and adjacent normal tissue samples from 72 OSCC-GB patients, as well as leukoplakia tissue from 25 patients with concurrent leukoplakia.

**Findings:** Our analysis revealed activated epithelial-mesenchymal transition, angiogenesis, and cell-cycle function in OSCC-GB. We found significant enhancement of glycolysis and reduction in oxidative phosphorylation, which are hallmarks of the Warburg effect. Immune profiling indicated enriched immune-related genes and cells in tumor tissues. We identified two distinct patient subtypes, one of which exhibited higher immune cell infiltration and showed potential for greater responsiveness to immune checkpoint inhibitors. *CD226*, *CD38*, and *KBTBD8* were identified as potential biomarkers for classifying OSCC-GB patients and were validated in an independent cohort. Significantly more M1 macrophages and CD4+ T-cells in leukoplakia tissue than the normal indicate activated host defense mechanisms in pre-malignant lesions, highlighting the potential for early intervention to prevent malignancy. TCGA-HNSC data exhibited similar gene set enrichments, including glycolysis and immune-related pathways. However, unique profiles in a subset of TCGA-HNSC patients highlight the molecular heterogeneity of head and neck cancers.

**Conclusion:** Our findings underscore the critical role of understanding these pathways in cancer biology and immunology, essential for developing effective treatment strategies for oral cancer and immunotherapy.

## Introduction

Globally, oral cancer is ranked sixteenth in incidence among all cancers ^1^. However, in India, the incidence of cancers of the oral cavity and the lip is the highest among males (∼16%) and the second highest when both sexes are considered (∼14%)^2^. The high incidence is attributable to widespread exposure to known environmental risk factors^3^, such as smokeless tobacco (chewing, dipping, and snuff), chewing betel quid containing areca nut, and alcohol consumption. Oral squamous cell carcinoma of the gingivo-buccal region (OSCC-GB) is a significant public health challenge in India^4^. Addressing this challenge requires prevention strategies and early detection efforts.

We have previously identified ^3^ recurrently-mutated driver genes for OSCC-GB, some of which are common to patients suffering from head and neck squamous cell carcinoma (HNSCC). The epigenomic landscape of OSCC-GB revealed significant dysregulation by epigenetic modification of DNA methylation-regulating genes ^4^. We found that extracellular matrix receptor interaction, focal adhesion, PI3K-Akt signaling, and cell cycle pathways were significantly enriched in OSCC-GB patients ^5^. Single-cell analysis revealed enrichment of partial epithelial-mesenchymal transition (EMT) in OSCC-GB malignant tumor populations ^6^. Significant promoter hypomethylation-driven upregulation of immune checkpoint ligands *PD-L1 (CD274)* and *CD80* was observed ^4^. Some genomic alterations in oral leukoplakia impact the immune dynamics of pre-cancer tissue, leading to immune suppression and progression to malignancy ^7^.

Head and neck cancers across anatomical sites are among the most highly immune-infiltrated, with diverse immune cells in the tumor microenvironment, including T cells, B cells, macrophages, and dendritic cells ^8^. The immune landscape is characterized by significant immune evasion mechanisms, such as the upregulation of immune checkpoint molecules *PD-L1 (CD274)* and *CTLA-4*, that inhibit T-cell function and allow tumors to escape immune surveillance ^9–12^. Additionally, gene alterations controlling the cell adhesion, EMT, and NF-κB signaling are observed in high-risk head and neck squamous cell carcinoma (HNSCC) ^13^; these alterations contribute to immunosuppression ^14^. Head and neck cancers also exhibit increased expression of cytokines and chemokines that attract immunosuppressive cells, further aiding tumor immune evasion ^8^.

Earlier studies on the transcriptomic profiles of OSCC-GB patients identified pathways significantly altered across all patients. Our study used a personalized approach by using single-sample gene set enrichment analysis with a paired tumor-normal design for each patient. We identified altered hallmark gene sets in the pooled patients and at the individual patient level. This approach enabled identification of immune-related differences among OSCC-GB patients and various distinguishing features based on other cellular functionalities. We also used the same methods on data of paired tumor-normal samples from head and neck cancer patients available for the TCGA cohort (TCGA-HNSC), comparing them with the anatomical subtype of OSCC-GB patients. We have identified distinct marker genes for the two OSCC-GB and the three TCGA-HNSC patient subtypes.

## Methods

### Patient recruitment and data acquisition

Our study cohort comprised 72 OSCC-GB patients enrolled at the Advanced Centre for Treatment, Research and Education in Cancer (ACTREC), Dr. R. Ahmed Dental College & Hospital (RADCH), and Chittaranjan National Cancer Institute (CNCI). Tumor and adjacent normal tissue samples were collected from each patient. We considered only those tumor samples with ≥ 80% of tumor cells reported on pathological reviews. Additionally, we collected histologically verified leukoplakia tissue samples from 25 of 72 OSCC-GB patients with associated oral leukoplakia. Written informed consent was obtained from every participant recruited in this study. The ACTREC, RADCH, CNCI, NIBMG, and ISI Review Boards approved the study. Clinical information and tissue samples of OSCC-GB patients recruited in our study are provided in Table S1 and Fig. 1A.

**Figure 1:**
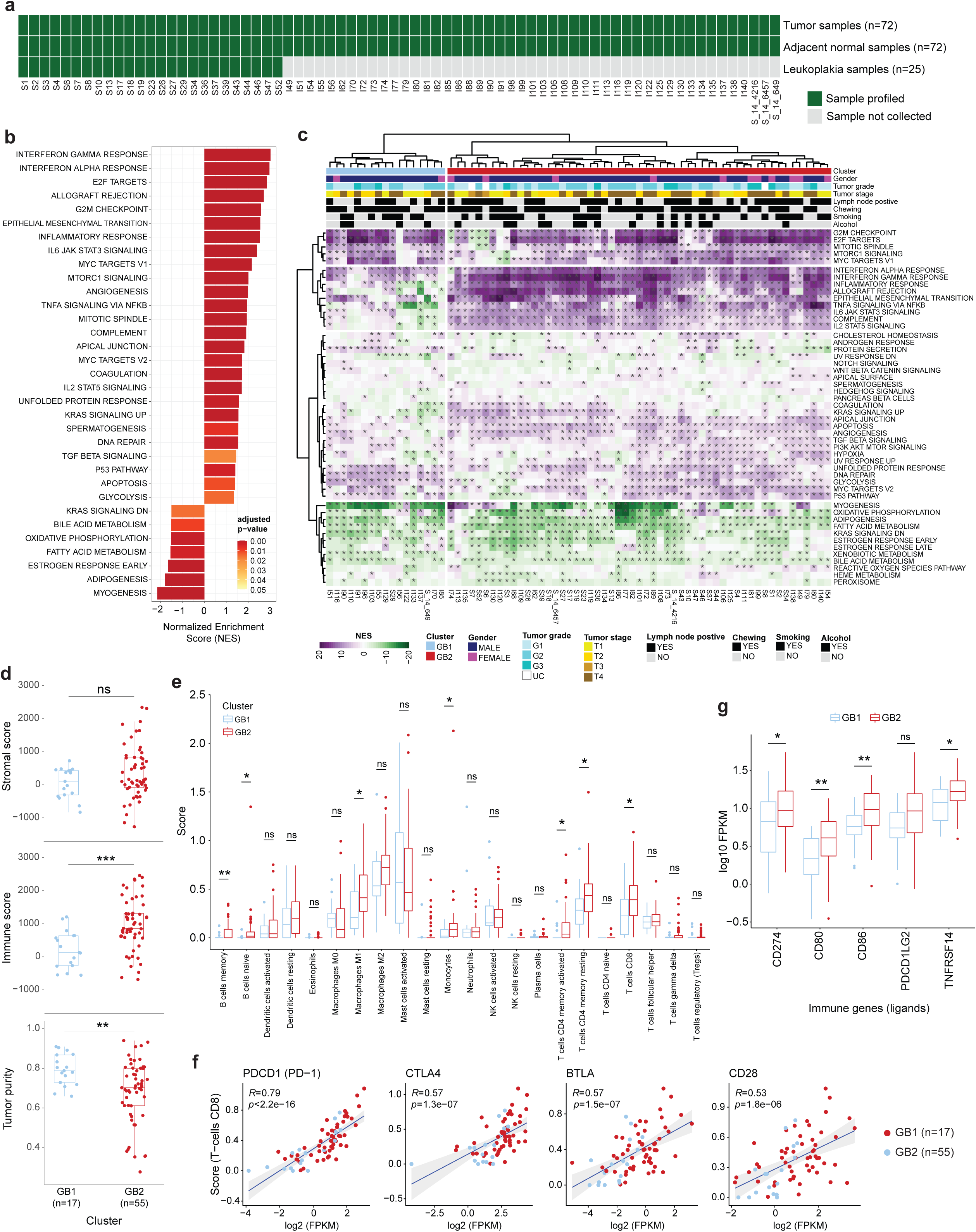
The transcriptomic landscape of OSCC-GB patients. (a) Data availability for the study participants. (b) Hallmark gene sets significantly enriched in OSCC-GB. (c) Unsupervised hierarchical clustering of OSCC-GB patients (n=72) using normalized enrichment scores of hallmark gene sets. Asterisks indicate significant (FDR < 0.05) enrichments. (d) Differential immune cell infiltration levels and tumor purity between GB clusters in OSCC-GB patients. (e) Comparison of 22 immune cell types abundances between GB1 and GB2 tumors of the OSCC-GB cohorts. (f) Scatter plots showing a significant positive correlation between abundance of CD8+ T-cells (CIBERSORT absolute) and the expression level (log2 FPKM) of four cell surface marker genes in OSCC-GB cohort of 72 patients. R values indicate Pearson’s correlation coefficients. (g) Expression differences of selected immune checkpoint ligands between the clusters. Statistical significance in d and g was determined using two-sided Mann-Whitney U tests. p-values for e were calculated using two-sided Welch’s t-tests. Significance level in d, e, and g: *, p<0.05; **, p<0.01; ***, p<0.001.

The gene expression profiles of 520 HNSCC patients from TCGA Pan-Cancer Atlas datasets were downloaded from the UCSC Xena (https://xena.ucsc.edu/) in October 2023. For further analyses, we considered 471 TCGA-HNSC patients with no evidence of metastasis. Adjacent normal samples were available for 41 of 471 TCGA-HNSC. Detailed clinical information for all patients was obtained from the TCGA Pan-Cancer Atlas (https://gdc.cancer.gov/about-data/publications/pancanatlas).

### RNA sequencing and data processing

Total RNA was extracted from all tissue samples of 72 OSCC-GB patients using AllPrep DNA/RNA Mini Kit (QIAGEN, Hilden, Germany). The concentration and quality of the isolated total RNA were assessed using a NanoDrop 2000 spectrophotometer and an Agilent 2100 Bioanalyzer with the RNA 6000 Nano Kit (Thermo Fisher Scientific, Waltham, MA). All samples had an OD260/OD280 ratio of ≥ 2 and an RNA Integrity Number (RIN) of ≥ 7.0. We used the TrueSeq RNA Sample Preparation Kit (Illumina, CA) to prepare sequencing cDNA libraries from the RNA samples following rRNA depletion. We assessed the quality and quantity of the cDNA libraries using an Agilent 2100 Bioanalyzer with a High Sensitivity DNA chip and a 7900HT Fast Real-Time PCR System (Life Technologies) with the Kapa Library Quantification Kit (Kapa Biosystems). We conducted 100 bp paired-end sequencing for each triplex cDNA library pool using the HiSeq-2000 and HiSeq-2500 platforms (Illumina, CA). All assays were carried out following the manufacturer’s protocols.

The quality of the FASTQ files, generated by demultiplexing the raw sequencing data, was assessed using FastQC (v. 0.10.1) software (www.bioinformatics.babraham.ac.uk/projects/fastqc/). Over 50 million high-quality paired-end RNA sequence reads were generated from each tumor, leukoplakia, and adjacent normal tissue sample. The paired-end RNA-seq reads were aligned to the human reference transcript (ftp://ftp.illumina.com/Homo_sapiens/UCSC/hg19/) and reference genome (hg19) using Tuxedo protocol [TopHat2 (v. 2.1.1) ^15^, Bowtie 2 (v. 2.3.4.1) ^16^], and SAMtools (v. 0.1.19) ^17^, with default settings. To reduce false positives, multi-mapped and non-concordant reads were discarded using SAMtools, and duplicate reads were identified and removed with PICARD’s MarkDuplicates (v. 2.17.0) tool (https://github.com/broadinstitute/picard). Mapped transcripts were assembled with Cufflinks (v. 2.2.1) ^18^ using default settings. Normalized gene expression (FPKM) values for all samples in the OSCC-GB cohort were calculated using Cuffnorm (v. 2.2.1) ^18^ with default parameters.

### Gene set enrichment analysis (GSEA) and single-sample GSEA (ssGSEA)

The list of expressed (average FPKM > 1) genes was ranked using average log2 (fold-change) in tumor compared to paired adjacent-normal samples from (a) 72 OSCC-GB and (b) 41 TCGA-HNSC. The ranked genes from OSCC-GB and TCGA-HNSC datasets were used to run the GSEA algorithm with R-based clusterProfiler (v3.18.1) ^19^ package for assessing the enrichment of hallmark gene set collections (n=50) of the Molecular Signatures Database (MSigDB v7.4) ^20^. All parameters were set at default for GSEA using clusterProfiler.

The list of genes used for GSEA analyses described above was ranked for each OSCC-GB (n=72) and TCGA-HNSC (n=41) using log2 (fold-change) between tumor and paired normal tissue samples. Single-sample gene set enrichment analysis (ssGSEA) to investigate the enrichment of hallmark gene sets (n=50) was performed with R package ssGSEA2.0 (https://github.com/broadinstitute/ssGSEA2.0), using default settings.

### Characterization of the immune microenvironment

We applied the ESTIMATE algorithm using the R-based estimate package (v. 1.0.13) ^21^ to identify the infiltration level of immune and stromal cells in each tumor from OSCC-GB (n=72) and TCGA-HNSC (n=471). CIBERSORT deconvolution algorithm ^22^ was used to characterize the immune cell compositions (n=22) from the gene expression profiles of all tumor, adjacent normal, and leukoplakia tissue samples. CIBERSORT was run in “absolute mode” using 1000 permutations to generate the p-values.

### Statistical analysis

Similar statistical analysis strategies were used for identifying biomarkers to classify OSCC-GB (n=72) or TCGA-HNSC (n=41) patient cohorts based on transcriptome profiles of their tumor samples. Partial least squares discriminant analysis (PLS-DA) algorithm ^23^ was applied independently on OSCC-GB and TCGA-HNSC to distinguish patient clusters within each cohort. For our PLS-DA models, we run R (Bioconductor) package ropls (v. 1.22.0) ^24^ using pareto-scaled gene expression (FPKM) of tumor samples. Variable importance in the projection (VIP) scores indicated the contribution of each gene to our models. We selected OSCC-GB or TCGA-HNSC patients. For downstream analyses on identifying biomarkers of OSCC-GB or TCGA-HNSC patients, we only considered genes with a VIP > 1 in each cohort. We then performed a t-test between tumor samples of two GB clusters of OSCC-GB patients.

Similarly, t-tests between tumor samples of each two of all three HNSCC clusters of TCGA-HNSC patient cohorts were done. We assessed the diagnostic performances of selected biomarkers (genes) for classifying (a) OSCC-GB to respective GB (n=2) and (b) TCGA-HNSC to respective HNSCC (n=3) clusters, using Leave-One-Out Cross-Validation (LOOCV) of logistic regression model ^25^.

### Validation of OSCC-GB biomarkers using external data

Microarray data for validating biomarkers were obtained from the Gene Expression Omnibus (GEO) repository (https://www.ncbi.nlm.nih.gov/geo/) with accession number GSE85195. The dataset was downloaded in its raw format for thorough preprocessing and analysis. The expression data were quantile-normalized using R (version 4.4.2) before conducting logistic regression-based prediction modeling. For single-sample Gene Set Enrichment Analysis (ssGSEA), genes were ranked based on log2 fold-change values calculated for each OSCC-GB tumor sample (n=34) compared to the matched normal control. The immune cell compositions within all tumor samples were characterized using CIBERSORT, which was run in “absolute mode” with 1,000 permutations.

## Results

### Patient cohorts and sample collection

We collected tumor and adjacent normal tissue samples from a cohort of 72 OSCC-GB patients. Additionally, leukoplakia tissue samples were collected from 25 of the 72 patients each of whom had concomitant leukoplakia in the oral cavity (Fig. 1a).

As expected, the majority of patients were male (81.9%) and habitual users of tobacco, primarily chewers (87.5%) many of whom (33.3%) were also smokers. The proportion of patients who presented with low (T1 + T2) or high (T3 + T4) tumor stages were nearly equal; 51.4% and 48.6%, respectively. Detailed demographic and histopathological characteristics of the study participants are provided in Table S1.

### Enriched hallmark gene sets identified two clusters of OSCC-GB patients

Gene set enrichment analysis (GSEA) identified 33 hallmark gene sets significantly (adjusted p-value < 0.05) enriched in tumor samples compared to paired normal samples (Fig. 1b). Positive enrichment of 78.8% gene sets indicated higher activation in tumor tissues compared to the normal of several immune functions (including interferon alpha and gamma responses; inflammatory response; IL-6/JAK2/STAT3 signaling), angiogenesis, cell-cycle-related functions (including E2F targets, G2M checkpoint, mitotic spindle), TGFβ signaling, p53 pathway and glycolysis. Overall, negative enrichments of gene sets related to metabolism (e.g., bile-acid and fatty-acid metabolisms, adipogenesis) and oxidative phosphorylation indicated their deactivation in OSCC-GB tumors (Fig. 1b).

Despite a uniform direction of significant dysregulation of gene sets in many tumors, some patients were found to have enrichment in the opposite direction. Cell-cycle-related gene sets were predominantly positively (NES>0) enriched, except for a few patients. MTORC1 signaling was significantly (FDR<0.05) positively enriched in ∼83% of OSCC-GB patients. Tumors in most OSCC-GB patients had significant (FDR<0.05) negative (NES<0) enrichment of gene sets related to fatty-acid metabolism (86.1%), adipogenesis (86.1%), and bile-acid metabolism (72.2%) compared with adjacent normal tissue samples.

Unsupervised clustering of the OSCC-GB cohort, using enrichment scores from hallmark gene sets (n=50), grouped the patients into two clusters (denoted as GB1 and GB2) (Fig. 1c). Compared to patients in GB1, among patients of GB2 there was significantly higher proportion of positively enriched gene sets related to apoptosis (94.5% vs. 23.5%), apical junction (76.4% vs. 35.3%), and angiogenesis (76.4% vs. 35.3%). The trend was the opposite for DNA repair (94.1% of GB1 vs. 58.2% of GB2) and glycolysis (94.1% of GB1 vs. 60% of GB2). All patients in GB2 showed enrichment of the hallmark IL-6/JAK/STAT3 signaling at significantly (FDR<0.05) higher levels in tumors than in normal samples, whereas ∼ 53% GB1 patients had negative enrichment scores (NESs).

To compare the similarity among the patients in respect of gene set enrichment, the cosine similarity between pairs of patients was measured using NES values from 50 hallmark gene sets. Intra-cluster similarity of patients was much higher than inter-cluster similarity; only a small set of GB1 patients were similar to patients of GB2 (Fig. S1). It may be noted that no significant differences were found (Fisher’s exact test, p>0.05) between GB1 and GB2 patients for any of clinical and histological features presented in Fig. 1c. Thus, genomic data are able to provide deeper insights into the extent and nature of similarity among OSCC-GB patients.

Our observation of significant (Mann-Whitney U test, p<0.0001) differential enrichments of immune-related hallmark gene sets between GB1 and GB2 patients (Fig. S2) prompted us to investigate the tumor immune microenvironment profiles in these two clusters of patients. We found significantly (Mann-Whitney U test, p<0.05) higher immune scores (p=0.0004) and lower tumor purities (p=0.006) in GB2 tumors than in GB1 (Fig. 1d). Stromal scores were roughly the same between the subtypes. We estimated the abundances of 22 immune cell types in the tumor samples using the CIBERSORT absolute deconvolution method. The CD4+ memory (activated/resting), CD8+ T-cells, B cells (memory/naive), M1 macrophages, and monocytes were significantly (t-test, p<0.05) more abundant in GB2 tumors than GB1 tumors (Fig. 1e).

Significant positive (Pearson correlation; p<0.05, R>0) association between expressions (log2 FPKM) of immune checkpoint receptors (ICRs) – *PDCD1* (*PD-1), CTLA4, BTLA, CD28* – and CD8+ T-cell abundances (CIBERSORT absolute) were observed in the tumor samples (Fig. 1f). These ICRs were also positively correlated with CD4+ memory T-cells (Fig. S3a, S3b). We did not find significant differences between patients of GB2 and GB1 clusters for genes belonging to the Human Major Histocompatibility Complex (MHC) or the Human Leukocyte Antigen (HLA) system (Fig. S4). *HLA-A/B/C* genes were expressed in tumors from both subtypes. We found significantly higher (Mann-Whitney U test, p<0.05) expression of immune checkpoint ligands (ICLs), including *CD274* (*PD-L1*), *CD80*, *CD86*, and *TNFRSF14* (*HVEM*), in GB2 tumors compared to GB1 tumors (Fig. 1g). Expressions of *PDCD1LG2* (*PD-L2*) and three other ICLs were also higher in GB2 than GB1 tumors, but not at statistically significant levels (Fig. 1g, S5). The significant (Pearson correlation, p<0.05) positive correlations observed between the expressions of ICRs and corresponding ICLs – *PD-1* vs. *PD-L1* (p=3.2×10^−6^) or *PD-L2* (p=1.1×10^−5^), *CTLA4* vs. *CD80* (p<2.2×10^−16^) or *CD86* (p<2.2×10^−16^), *BTLA* vs. *HVEM* (p=5.6×10^−14^), and *CD28* vs. *CD80* (p=3.6×10^−10^) and *CD86* (p=2.2×10^−14^) – in the OSCC-GB tumors (Fig. S6) suggests an active immune checkpoint blockade mechanism. Together, our results indicate patients of the GB2 subtype are immunologically hot with higher T-cell infiltrations in tumor tissues. Thus, GB2 patients may respond better to immune checkpoint inhibition therapy than their GB1 counterparts.

### Progressive upregulation of immune checkpoint ligands from normal through leukoplakia to tumor tissues

We explored the trends of infiltrated immune cells (n=6) from normal tissue to leukoplakia to tumor among patients belonging to GB2 (n=24). We found significantly (p<0.05) higher infiltration of M1 macrophages in (a) tumor compared to paired leukoplakia tissues and (b) leukoplakia tissues compared to paired normal tissues (Fig. 2a). Memory B-cells, and memory CD4+ T-cells (activated/resting) were significantly more infiltrated in tumor and leukoplakia tissues than adjacent normal oral tissue samples; no significant differences were observed between tumor and leukoplakia samples. Statistically significant differences were not observed between tumor and adjacent normal samples of for the three immune cell types – CD8+ T-cells, naïve B-cells, and monocytes (Fig. 2b). Though we failed to find significant differences in the infiltration rates, as evidenced by higher CIBERSORT scores, of these immune cells (except for M1 macrophages), monotonic dysregulation of ICLs including *PD-L1*, *PD-L2*, *CD86*, and *CD86* was observed during progression from normal through leukoplakia to tumor tissues of OSCC-GB patients (fig. 2c). ICRs like *PD-1, CTLA4*, and *BTLA*, were significantly (Pearson correlation, p<0.05) positively correlated both with CD8+, and CD4+ T-cells (Fig. S7). Thus, for GB2 patients, the drugs targeting PD-1 or CTLA4 may be equally effective in treating oral cancer or leukoplakia (oral pre-cancer). The combination of PD-L1 and CD80/CD86 inhibitors could be helpful for the treatment of GB2 patients or individuals with leukoplakia.

**Figure 2:**
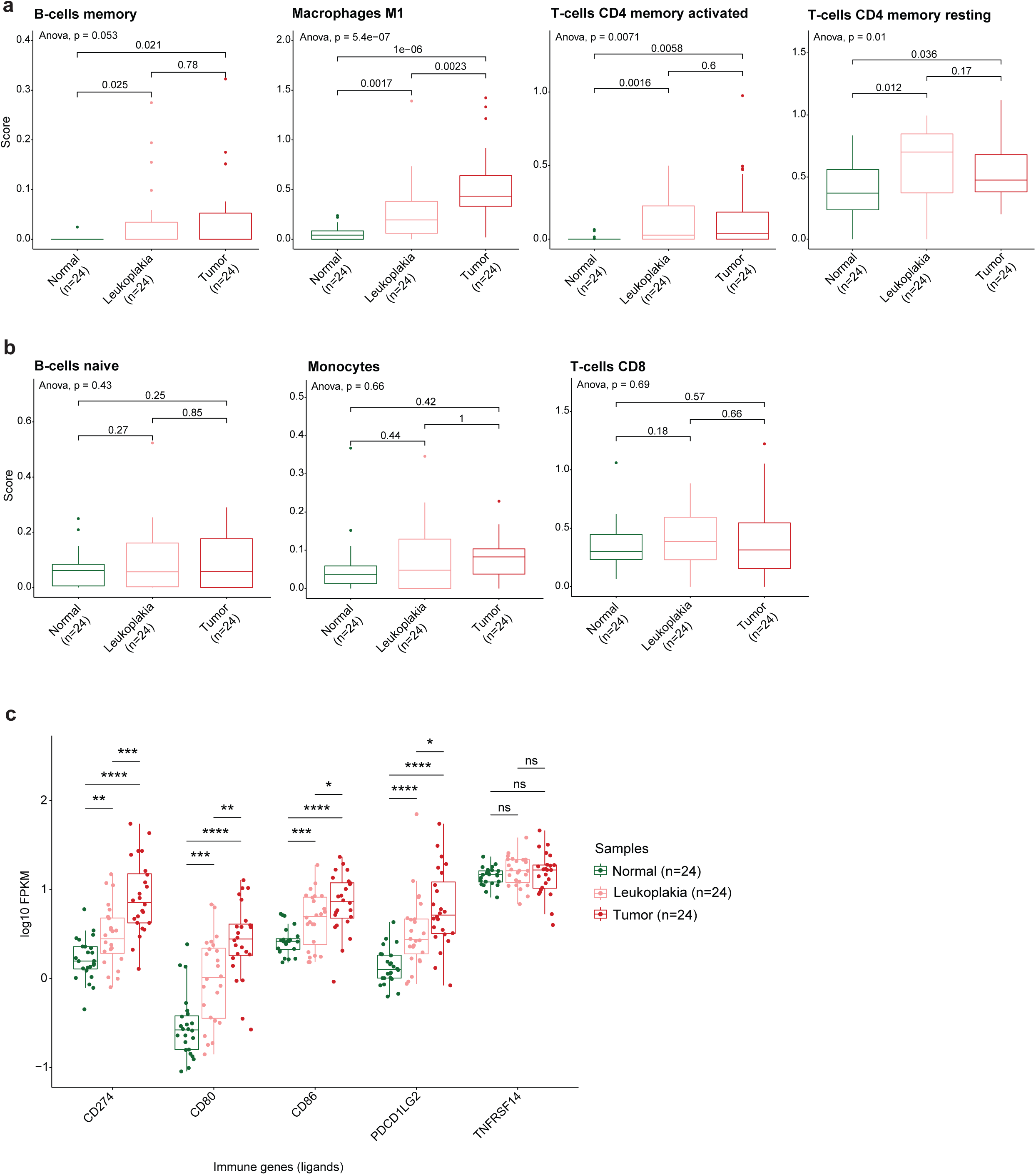
Leukoplakia tissues with immune microenvironment similar to tumors but with intermediate expression of immune ligands between normal and tumor samples. (a) Plots showing immune cells in leukoplakia tissues that infiltrated at significantly higher levels than normal but similar to tumor tissues. (b) Plots indicating similar infiltration patterns of immune cells in normal-leukoplakia-tumor triad tissue samples. For a-b, p-values between any two pairs were calculated using two-sided paired t-tests. (c) Monotonic upregulation of immune ligands in normal, leukoplakia, and tumor tissues. Statistical significance was determined using a two-sided Wilcoxon signed-rank test. Significance level: *, p<0.05; **, p<0.01; ***, p<0.001; ****, p<0.0001.

### Gene set enrichment analysis identifies TCGA-HNSC subtypes with different enrichments of hallmark gene sets

We used the GSEA algorithm to identify enrichment of hallmark gene sets in 41 TCGA-HNSC patients with paired tumor-normal data. In tumors, 38 gene sets were significantly enriched (adjusted p-value < 0.05), with 74% positively enrichment. The most enriched gene sets were related to positive enrichments of cell-cycle, immune system, angiogenesis, EMT pathways, or negative enrichments of metabolism, adipogenesis, oxidative phosphorylation, and myogenesis pathways (Fig. 3a).

**Figure 3:**
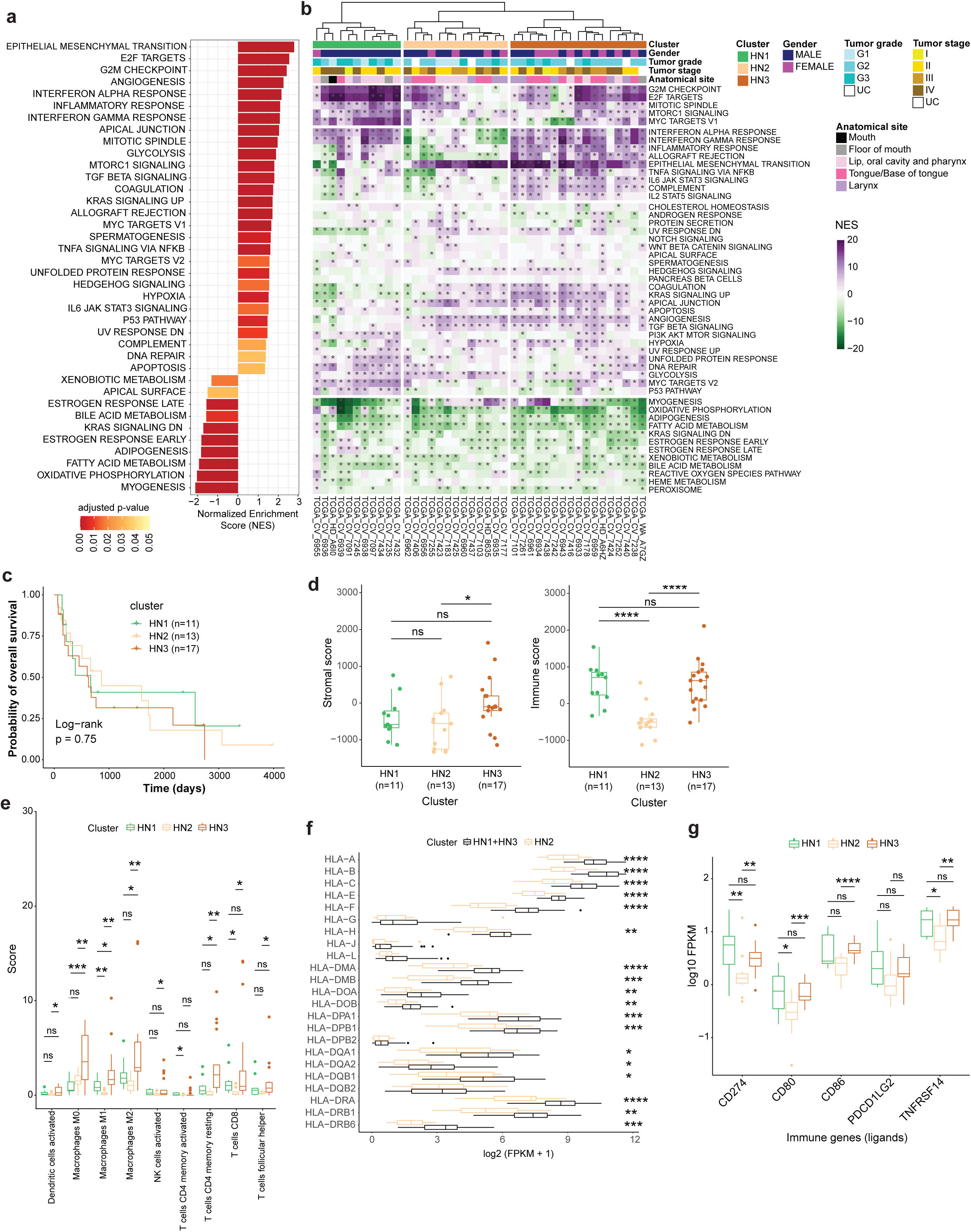
The transcriptomic landscape of TCGA-HNSC patients for whom paired tumor-normal tissues were available. (a) Significantly enriched hallmark gene sets in the TCGA-HNSC cohort. (b) Unsupervised hierarchical clustering of TCGA-HNSC patients (n=41) using activities of hallmark gene sets. The gene set activities are based on NES from ssGSEA analysis. Asterisks indicate significant (FDR < 0.05) enrichments. (c) Kaplan-Meier curve for overall survival in the TCGA-HNSC cohort stratified by three head and neck (HN) clusters – HN1, HN2, and HN3. (d) Differential infiltrations of immune and stromal cells in tumors of HN patient clusters. (e) Abundances of immune cell types that significantly differed between one or more pair(s) of HN clusters in the TCGA-HNSC cohort. (f) Significantly higher expression of most HLA genes observed in immunologically hot (HN1 and HN3) compared to cold (HN2) tumors. (g) Plots showing consistently low expression of immune ligands in HN2 than in other patient clusters. Statistical significance in d, f, and g was determined using two-sided Mann-Whitney U tests. For e, p-values were calculated using two-sided Welch’s t-tests. Significance level in d-g: *, p<0.05; **, p<0.01; ***, p<0.001; ****, p<0.0001.

We performed unsupervised clustering of HNSCC (n=41) patients recruited in TCGA, using ssGSEA enrichment scores of hallmark gene sets. The patients were grouped into three clusters – HN1, HN2, and HN3 (Fig. 3b). Immune-related gene sets were the most positively enriched in HN3 and negatively enriched in HN2 clusters of patients (Fig. S8). The highest and lowest proportions of significant negative enrichments of the hallmark myogenesis gene set were seen in HN1 (100%) and HN2 (23%) clusters of patients, respectively. Myogenesis was mostly (∼61.5%) significantly (FDR<0.05) positively enriched in tumor than in normal samples of HN2 patients. Significant (FDR<0.05) positive EMT enrichments were seen in ∼92.3% HN2 and ∼27.3% HN1 patients. There were no tumors in HN2 patients, but about 36.4% of tumors in HN1 patients were significantly negatively enriched for EMT. As expected, high levels of similarity in the patterns of enrichment of hallmark gene sets were observed in HNSCC patients within each subtype (Fig. S9). There were notable differences among patients belonging to different subtypes. No significant (Log-rank test, p>0.05) difference in overall survival (Fig. 3c) or progression-free survival (Fig. S10) was observed among the subtypes.

We examined TCGA-HNSC tumors (n=41) using the ESTIMATE algorithm to identify immune and stromal scores. Significantly higher scores were observed in HN3 for stromal (p=0.012) and immune (p=2.62×10^−5^) than in HN2 (Fig. 3d). Nine of twenty-two immune cells, including CD8+, CD4+, and follicular helper T-cells, showed significant (t-test, p<0.05) differential abundance between one or more head and neck (HN) subtypes (Fig. 3e, S11). The expressions of the HLA (or MHC) genes were generally the lowest among HN2 patients. Similar trends were observed between HN1 and HN3 tumors (Fig. S12). Compared to HN2, the HN1 and HN3 patients together showed significantly (Mann-Whitney U test, p<0.05) higher expressions of HLA genes (Fig. 3f). The ICLs – *CD274* (*PD-L1*), *CD80*, *CD86*, and *TNFRSF14* (*HVEM*) – were significantly (Mann-Whitney U test, p<0.05) upregulated in HN3 compared to HN2 set of TCGA-HNSC tumors (Fig. 3g). There were no significant differences between HN1 and HN3 tumors for expressions of the ICLs (Fig. 3g, S13). We observed significantly high positive correlations between the ICR-ICL pairs, including *PD-1* vs. *PD-L1* (p=7.5×10^−6^) and *PD-L2* (p=8×10^−4^), *CTLA4* vs. *CD80* (p=1.6×10^−7^) and *CD86* (p=3.5×10^−7^), *BTLA* vs. *HVEM* (p=1.9×10^−6^) in the tumor samples of TCGA-HNSC patients (Fig. S14), indicating an active immune checkpoint blockade mechanism. Thus, HN1 and HN3 subtypes appear as immunologically hot and HN2 as immunologically cold HNSCC patients.

### Transcriptome-wide comparison between squamous cell carcinomas of gingivo-buccal and other head and neck regions revealed large similarities, but also notable differences

We found an overall significant (adjusted p-value<0.05) enrichment of 39 hallmark gene sets among patients of either OSCC-GB or TCGA-HNSC, or both (Fig. 4a). Of the 39 gene sets, 32 (=25 positive + 7 negative) were enriched in both patient cohorts. The IL2–STAT5 signaling was significantly enriched in OSCC-GB patients. Among TCGA-HNSC patients, the gene sets were significantly positively enriched for hedgehog signaling and UV response DN (down), and negatively for apical surface. However, opposite enrichment patterns of these gene sets, though statistically non-significant (adjusted p-value>0.05), were observed in OSCC-GB patients.

**Figure 4:**
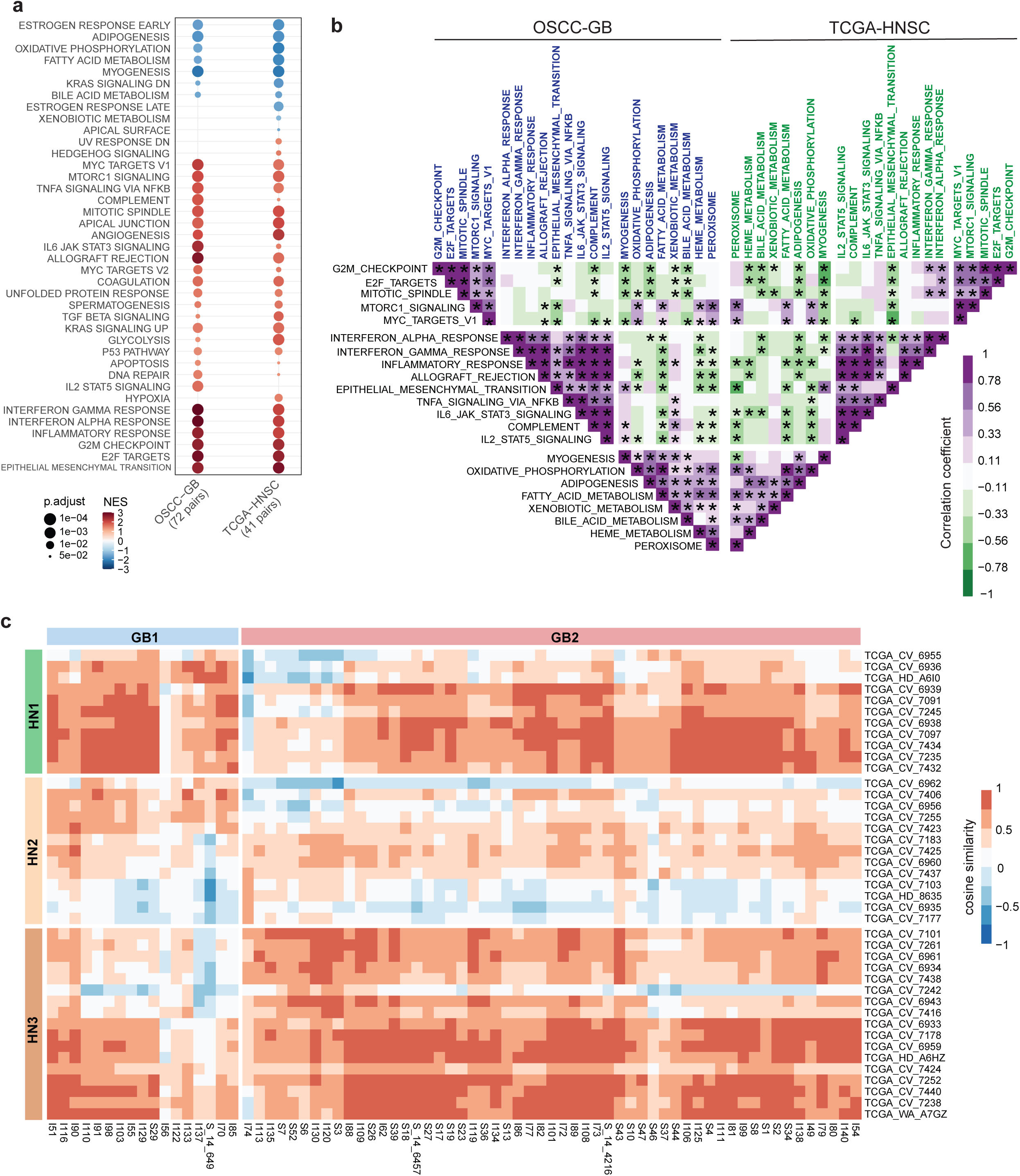
Comparison between transcriptomic profiles of OSCC-GB and TCGA-HNSC cohorts. (a) Significantly enriched hallmark gene sets in both OSCC-GB and TCGA-HNSC cohorts reveal similar enrichment patterns. Red dots, positive enrichments. Blue dots, negative enrichments. (b) Enrichment correlations of cell-cycle-, immune-, and selected metabolism-related hallmark gene sets using NES from 72 OSCC-GB (gene sets marked blue) or 41 TCGA-HNSC (gene sets marked green) cohorts. The color of the matrix represents Pearson’s correlation coefficients. An asterisk indicates significant (p-value < 0.05) correlation. (c) Heatmap illustrating the similarities between patients from GB clusters and those from HN clusters. The cosine distance metric calculated based on enrichment scores of hallmark gene sets was used to estimate similarities among patients from two cohorts.

We calculated the pairwise correlations of enrichment scores (NES) between gene sets from cell-cycle-, immune-, and metabolism-related hallmark gene sets for (a) OSCC-GB (n=72) and (b) TCGA-HNSC (n=41) patients (Fig. 4b). Irrespective of the patient cohort, cell-cycle related gene sets were significantly (p<0.05) positively correlated with each other. Positive correlations were also observed for immune– and metabolism-related gene sets. Among the TCGA-HNSC patients, interferon alpha response and myogenesis were significantly negatively correlated with epithelial-mesenchymal transition (p= 0.022, R=-0.36) and peroxisome (p= 0.0039, R= –0.44), respectively. These correlation coefficients were, however, positive (i.e., interferon alpha response vs. EMT, R=0.15; and myogenesis vs. peroxisome, R=0.13) among the OSCC-GB patients. The proportion of positive correlation coefficients between functionally similar gene sets was greater than that between functionally different gene sets, in both the patient cohorts (Fig. 4b).

While comparing the OSCC-GB patients with the TCGA-HNSC patients, we found GB1 and GB2 patient subtypes were similar (cosine similarity>0) to HN1 and HN3 patients, respectively (Fig. 4c). But, the head and neck cancer (TCGA-HNSC) patients from HN2 subtype were distinct from GB patients – they have the least (either negative or low positive) cosine similarities with both GB patient subtypes. Thus, we can infer that although the OSCC-GB and TCGA-HNSC patients displayed similar profiles, a proportion of patients from the latter cohort turned out to be different.

### Identification of biomarkers specific to gingivo-buccal or head and neck cancers

We applied a partial least squares discriminant analysis (PLS-DA) algorithm to distinguish OSCC-GB tumor subtypes (Fig. 5a). About 33% of the genes (=4431 of 13413) were detected with a variable importance in the projection (VIP) > 1 (Table S2). We performed a t-test on log2 transformed FPKM values of these genes for identifying differential expressions between GB1 (n=17) and GB2 (n=55) tumors. We found that 235 genes were significantly (Benjamini-Hochberg corrected p-value<0.01) differentially expressed by ≥2-fold in GB1 compared to GB2 tumor samples (Table S3). We selected three (∼1% of 235) most significantly differentially expressed genes as potential biomarkers for classifying OSCC-GB patients. The average accuracy of the model to classify GB1 and GB2 tumors using log2 (FPKM) of *CD226*, *CD38*, and *KBTBD8* was 86.1% (estimated using the leave-one-out cross-validation (LOOCV) method of logistic regression). The accuracy of correct classification using these three genes was much lower (53.6%) for HN1 (n=11) and HN3 (n=17) tumors. Thus, the biomarkers that distinguish GB1 and GB2 subsets of OSCC-GB patients rather well, perform poorly to distinguish between HN1 and HN3 subtypes of HNSCC patients. This indicates that different sets of biomarkers may be required for identifying subtypes of patients in distinct geographical regions.

**Figure 5:**
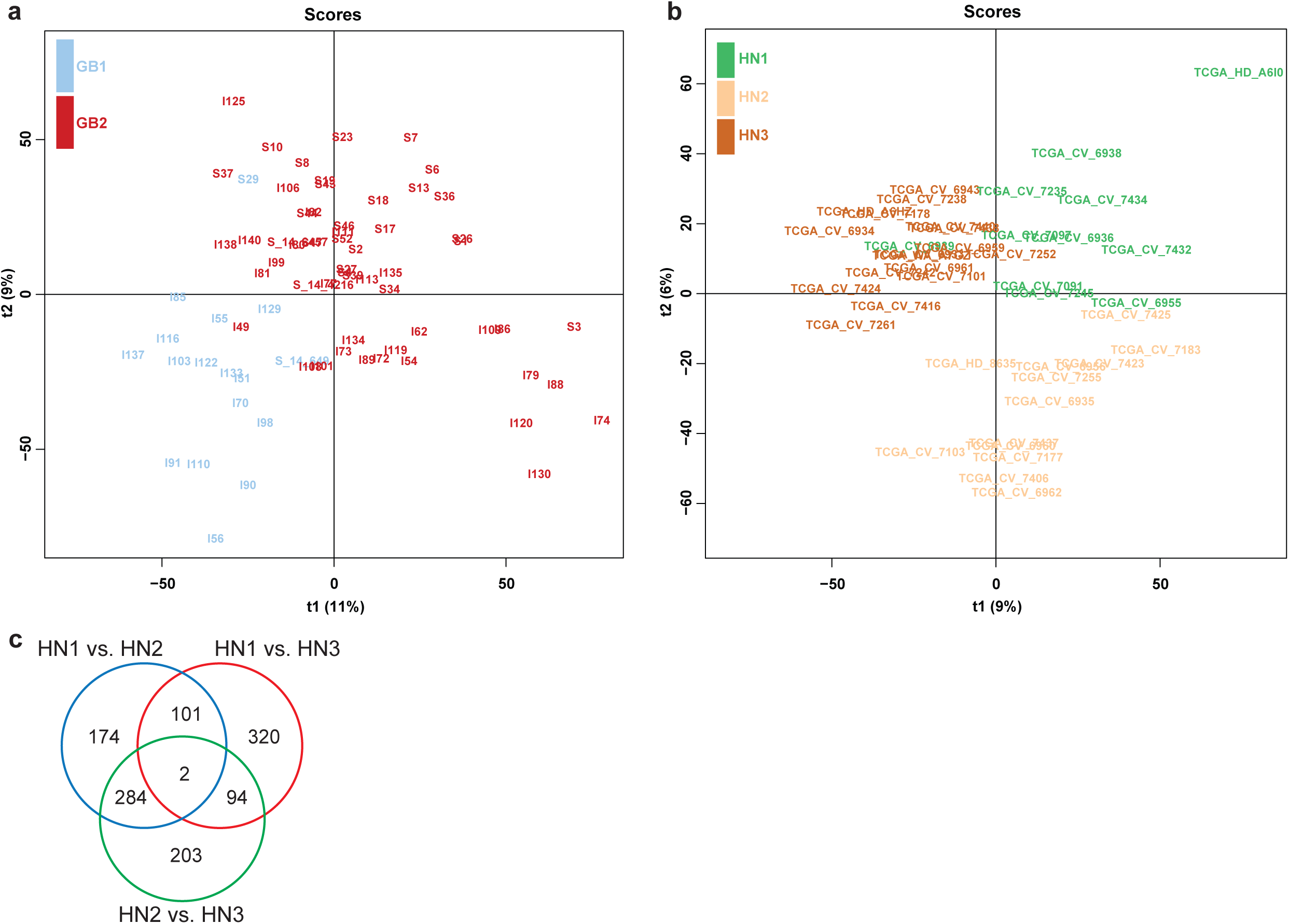
Identifying specific biomarkers to accurately predict OSCC-GB or HNSC tumor clusters. PLS-DA score plots show the segregation of patients from (a) OSCC-GB and (b) TCGA-HNSC cohorts. (c) Venn diagram representing the numbers of significantly differentially expressed genes between tumor samples of any pair of HN clusters.

We identified the expression levels (FPKM) of 6028 (of 15070) genes with VIP > 1 (Table S4) from the OPLS-DA model of TCGA-HNSC patients (Fig. 5b). We tested the equality of mean log_2_FPKM values of these genes between pairs of patient clusters [HN1 (n=11) vs. HN2 (n=13); HN1 (n=11) vs. HN3 (n=17); and HN2 (n=13) vs. HN3 (n=17)] and found that 8.5 – 9.7% of the genes were significantly (Benjamini Hochberg corrected p-value < 0.05) differentially expressed by ≥2-fold change between two HN tumor subtypes (Table S5-S7). *LPXN* and *ETV7* were significantly differentially expressed in all pairwise comparisons (Fig. 5c). The predictive ability (as determined by the average accuracy of the LOOCV of the logistic regression model) of the expression levels (log2 FPKM) of these genes as classifiers of subgroups of HN tumors were for HN1 (n=11) = 0.854, for HN2 (n=13) = 0.902 and for HN3 (n=17) = 0.732, respectively. Using the consensus of three predicted outcomes, we categorized 234 patients into one of three HN subtypes. Of 234 TCGA-HNSC patients, 44 (19%), 15 (6%), and 175 (75%) were classified as HN1, HN2, and HN3 subtypes, respectively (Table S9).

### Predicting cluster association for GSE85195 using OSCC-GB

We applied logistic regression to predict the cluster association for GSE85195 patients (n=34) using the normalized expression levels of *CD226*, *CD38*, and *KBTBD8* from tumor samples. The model was trained with log2 FPKM values from 72 OSCC-GB tumor samples as test data. All GSE85195 patients were predicted with high confidence (probability range: 0.99–1.00) to belong to the GB1 cluster.

Principal component analysis (PCA; Fig. 6a), based on NES of ssGSEA results from OSCC-GB and GSE85195, showed that GSE85195 samples clustered closely with GB1 OSCC-GB patients, highlighting their similarity to the GB1 subset. Additionally, a comparison of NES values for seven selected immune-related hallmark gene sets (Fig. 6b) revealed significantly lower levels in GSE85195 compared to the GB2 group of OSCC-GB patients.

**Figure 6:**
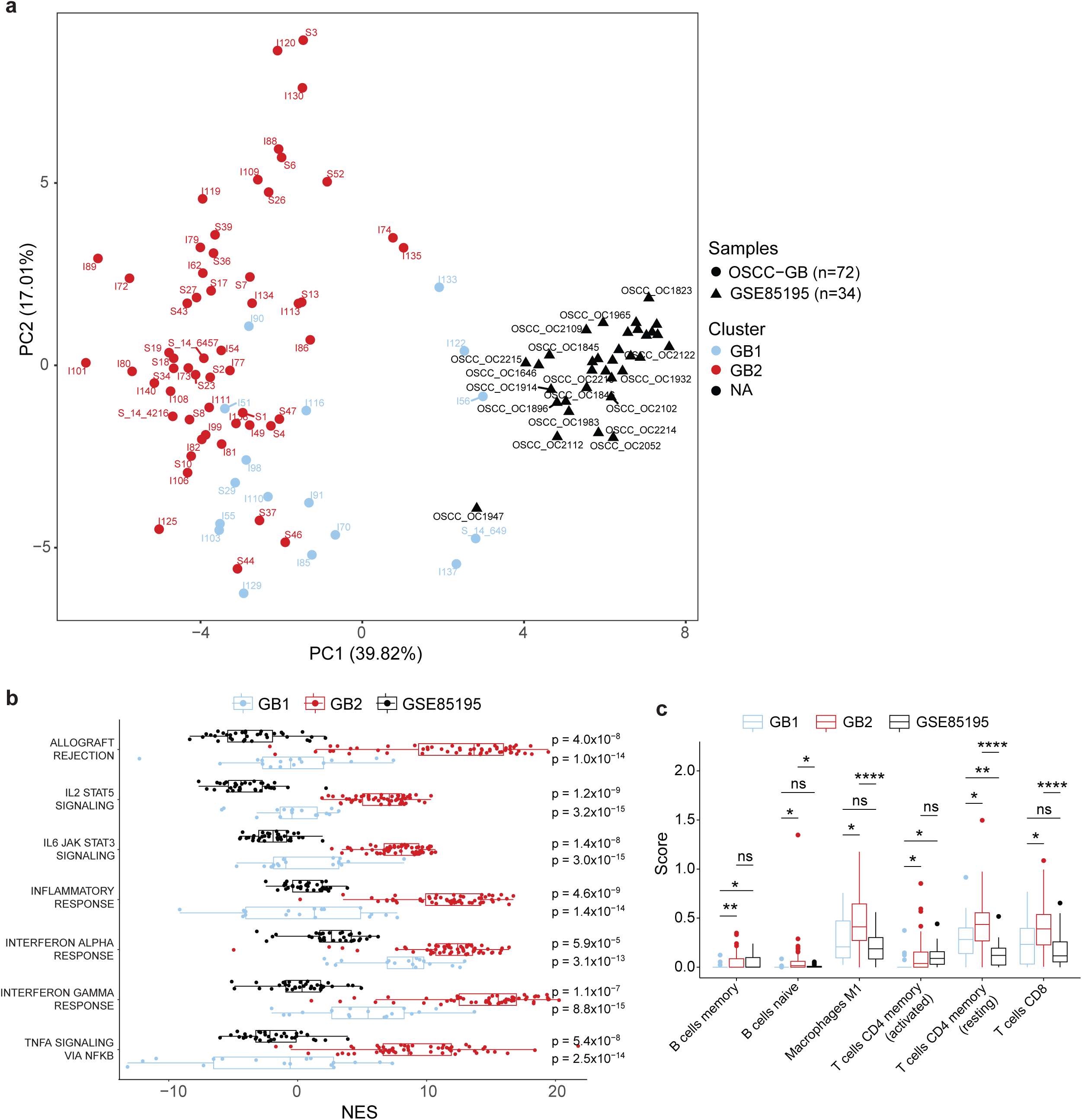
Cluster Association and Immune Profile Comparison of GSE85195 with OSCC-GB Patient Subgroups. (a) Principal component analysis of enrichment scores (NES) showing GSE85195 samples clustering closely with the GB1 OSCC-GB patient subgroup. (b) The comparison of NES for seven immune-related hallmark gene sets highlights significantly lower levels in GSE85195 and GB1 compared to the GB2 subgroup. (c) Infiltration levels of naive B cells, M1 macrophages, CD4+ (memory resting), and CD8+ T-cells are noticeably lower in GSE85195 and GB1 patients than in the GB2 set of OSCC-GB patients.

Furthermore, we examined the infiltration of six immune cell types that were significantly different between GB1 and GB2 patients (Fig. 1e), comparing these values against GSE85195 (Fig. 6c). Like GB1, GSE85195 patients showed significantly lower levels of naive B cells, M1 macrophages, CD4+ T-cells (memory resting), and CD8+ T-cells compared to the GB2 patient group.

### Predicted TCGA-HNSC tumor subtypes have similar immune profiles per the ssGSEA-based original clusters

We found significantly higher (Mann-Whitney U test, p < 0.05) immune (p=2.06×10^−6^) and stromal scores (p=4.33×10^−7^) in HN3 than HN2 tumors and significantly higher immune score (p=0.003) in HN1 compared to HN2 tumors (Fig. 7a). Patients with T3 and T4 tumor stages were significantly more prevalent (Fisher’s exact test, p= 0.034) in HN2 patients than in HN1 and HN3 patients combined. Though no statistically significant difference (log-rank test, p=0.2) in overall survival was noted between subtypes, the HN2 patients had the most poor prognosis compared to the other subtypes (Fig. 7b). A Multivariate Cox regression analysis was conducted to determine the impact of HN clusters on progression-free survival while controlling for age, gender, tumor grade, and risk factors, including tobacco smoking, alcohol consumption, and Human Papillomavirus (HPV) infections (Fig. S15). However, we could not establish any significant effect on progression-free survival in HN2 patients compared to HN1 and HN3 patients (Fig. S15). On profiling the TCGA-HNSC tumor samples for estimating the abundance of 22 immune cell types, we found significantly higher (Welch’s t-test, p<0.05) infiltrations of all macrophages (i.e., M0, M1, and M2), activated NK cells, CD4+ (memory resting), CD8+, and follicular helper T-cells in HN3 than HN2 tumors (Fig. 7c, S16).

**Figure 7:**
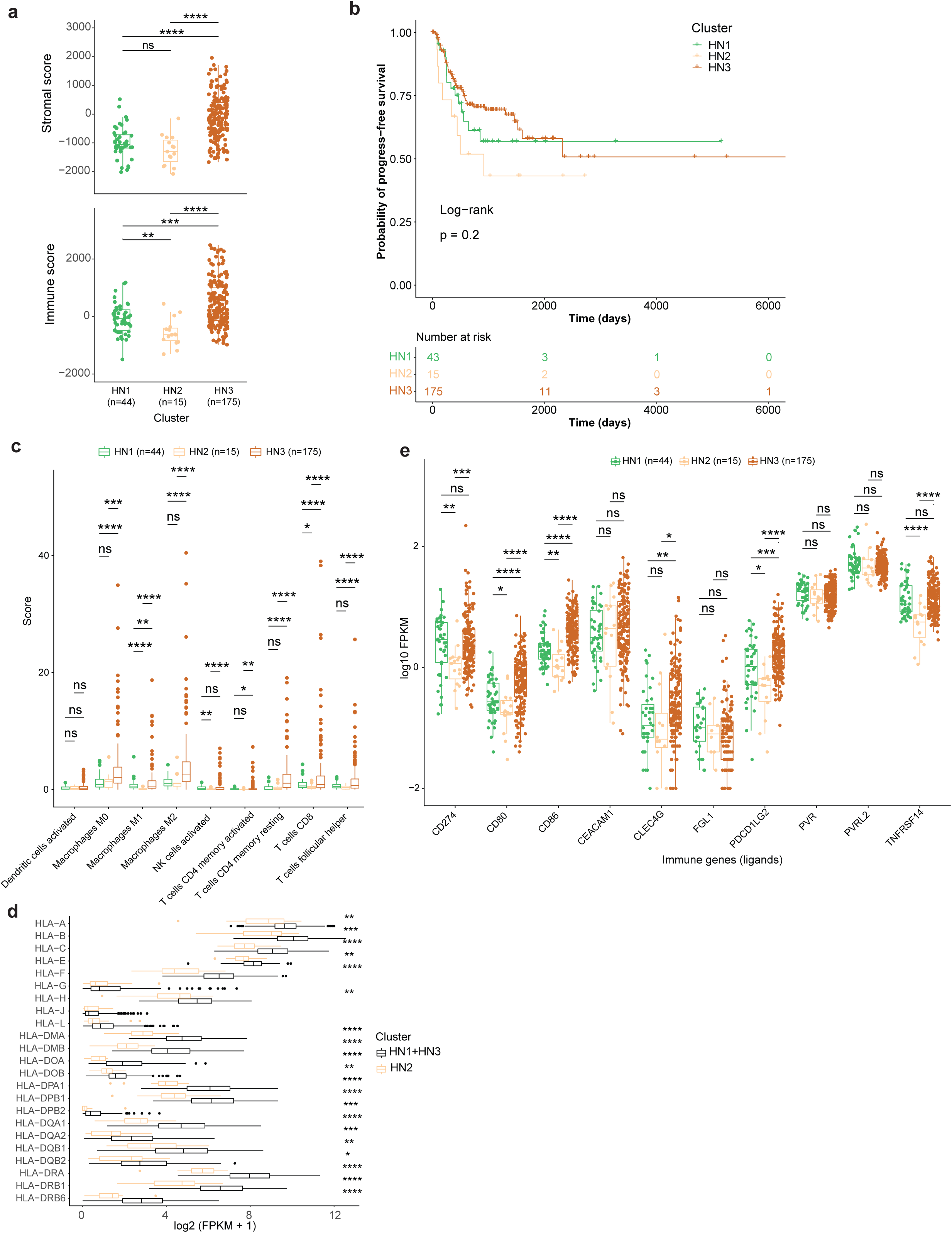
Immune landscapes of predicted clusters of TCGA-HNSC tumors. (a) Comparison of the stromal and immune scores between HN clusters in predicted TCGA-HNSC (n=234) cohorts. (b) Kaplan-Meier curve and the at-risk table for progression-free survival in predicted clusters of TCGA-HNSC patients. (c) Differential abundances of selected types of immune cells in patients from predicted HN clusters. (d) Comparison of HLA gene expressions between the immunologically hot (HN1 and HN3) and cold (HN2) tumors. (e) Differential expression of immune checkpoint ligands between the predicted clusters of TCGA-HNSC patients. Statistical significance in a, d, and e was determined using two-sided Mann-Whitney U tests. p-values for c were calculated using two-sided Welch’s t-tests. Significance level in a, c-e: *, p<0.05; **, p<0.01; ***, p<0.001; ****, p<0.0001.

Considering the relative expressions of HLA genes, the majority were significantly upregulated (Mann-Whitney U test, p<0.05) in tumors of HN1 and HN3 patients together (n=219) compared to that of HN2 patients (n=15) alone (Fig. 7d). We observed significant (Mann-Whitney U test, p<0.05) upregulations of several ICLs including *CD274* (*PD-L1*), *PDCD1LG2* (*PD-L1), CD80*, *CD86*, and *TNFRSF14* (*HVEM*) in both HN1 and HN3 compared to HN2 patient subtypes (Fig. 7e). No significant difference between HN1 and HN3 tumors was observed for *PD-L1* and *HVEM* expressions. ICRs, including *PDCD1 (PD-1)*, *CTLA4*, and *BTLA*, were found to significantly positively correlated (p<0.05) with CD8+ (Fig. S17a) and CD4+ T-cells (Fig. S17b, S17c). We observed a significant positive correlation (p<0.05) between the expressions (log2 FPKM) of some IC receptors and corresponding ligands, including *PD-1/PD-L1, PD-1/PD-L2, CTLA4/CD80, CTLA4/CD86*, and *BTLA/HVEM* in 234 TCGA-HNSC predicted subtypes (Fig. S18).

## Discussion

Our analysis of transcriptome data of tumor and adjacent normal samples from 72 patients has provided a comprehensive characterization of genes and pathways significantly and consistently altered in OSCC-GB. Our gene set enrichment analysis of OSCC-GB has revealed, consistent with some earlier findings, the activation of epithelial-mesenchymal transition (EMT) and angiogenesis ^26,27^, cell cycle-related functions ^28^ and significant positive enrichment of glycolysis and negative enrichment of oxidative phosphorylation in tumors compared to normal tissues, indicating the presence of the Warburg effect in the hypoxic tumor microenvironment ^29^.Further, our observation of altered fatty acid metabolism is indicative of sustained ATP production and macromolecular requirements for the growth of cancer cells ^30^. Our findings also suggest that the synthesis, decomposition, and transport of fatty acids are involved in various immune cell functions – destruction, activation, and differentiation – in the immune microenvironment of tumor cells ^31^. We observed significant positive enrichments of major immune function-related gene sets in tumors compared to adjacent normal samples of OSCC-GB patients. The expression of immune-related genes was upregulated as cells transformed to moderate dysplastic stages in a mouse OSCC model ^32^.

Immune landscape-based oral squamous cell carcinoma (OSCC) studies have identified immune-hot and immune-cold patient subtypes ^9^. Despite overall positive enrichment, significantly high enrichment of immune-related gene sets and higher immune scores in a subset of OSCC-GB patients (GB2 compared to GB1 patients) similarly indicates non-uniform immune involvement among OSCC-GB patients. Considerable enrichment of anti-tumor immune cells, including B-cells, M1 macrophages, CD4+, and CD8+ T-cells in the tumor microenvironments of GB2 patients suggests that those tumors were likely suppressed by host immune surveillance ^33^. Significantly high M1 macrophages and cytotoxic T-cell infiltrations in tumor microenvironments signify elevated pro-inflammatory and tumor-inhibitory functions among GB2 compared to GB1 patients. Classical major MHC I molecules were equally expressed in both GB1 and GB2. However, significant upregulation of ICLs, including PD-L1, CD80, CD86, and HVEM in GB2 compared to GB1 tumors, and a strong positive correlation with corresponding ICRs suggests that the GB2 subtype of patients is likely to be more responsive to immune checkpoint inhibition therapy compared to the other patients. BTLA/HVEM may be a potential target for treating GB2 tumors with checkpoint inhibition ^34^.

Our additional analysis of the transcriptome profiles of leukoplakia (pre-cancer) tissue samples collected from a subset of 25 OSCC-GB patients showed a higher density of M1 macrophages and CD4+ T-cells in microenvironments of leukoplakia than of adjacent normal samples, indicating that host defense mechanisms are activated even in pre-malignant lesions. Unlike higher abundance of CD8+ T-cells in proliferative leukoplakia compared to localized leukoplakia ^35^, we were unable to find significant changes in expression of CD8+ T-cells in immune-enriched GB2 subtype of OSCC-GB tumors and leukoplakia compared with adjacent normal tissues. Progressively high expression of ICLs, including PD-L1, PD-L2, CD80, and CD86, in leukoplakia than normal tissues indicate that pre-cancer tissues may be responsive to anti-PD-1 and anti-CTLA-4 combination therapy. We, however, note that an earlier report ^36^ indicated that proliferative verrucous leukoplakia tissues harboring 9p21.3 somatic copy-number loss developed OSCC even after treatment with anti-PD-1 drug nivolumab.

The TCGA-HNSC data indicate that HNSCC tumors exhibit significant enrichment of gene sets related to epithelial-mesenchymal transition (EMT) ^13^ and gene alterations regulating the cell cycle ^37^. Similar to OSCC-GB, there are positive enrichments of hallmark glycolysis, indicating the presence of Warburg effect in tumors of TCGA-HNSC patients. A previous study ^38^ on expression profiles of 520 TCGA-HNSC tumor samples revealed increased hallmark interferon-alpha response, myc targets, unfolded protein response, transforming growth factor-β (TGF-β) signaling, and interleukin 6-Janus kinase-signal transducer and activator of transcription 3 (IL-6/JAK/STAT3) signaling pathways in patients with upregulated glycolysis. Immune infiltration in TCGA-HNSC tumors, irrespective of HPV infections, suggests a complex interaction between the tumor and the immune system, particularly in HN1 and HN3 subtypes ^39^. Additionally, potential immune evasion mechanisms were observed at the single-cell level of HNSCC tumors during early metastasis ^40^. These findings underscore the critical role of understanding these pathways in cancer biology and immunology, as they are crucial in tumor progression, immune evasion, and potential therapeutic targeting for future treatment of HNSCC patients.

We identified three marker genes that correctly differentiate GB1 from GB2 tumors. Studies have shown optimal activation or re-activation of CD226 — expressed on the surface of natural killer (NK) cells and CD8+ T-cells — may be helpful as an adjunct to current PD-1/PD-L1 blockade treatments ^41^. CD38 inhibits CD8+ T-cell function via adenosine receptor signaling, and their blockade could be considered an effective strategy for overcoming resistance to PD-1/PD-L1 blockade ^42^. KBTBD8 controls the stability of proteins that regulate the cell cycle and cellular differentiation, playing a role in the proliferation and differentiation of various cell types ^43^. Together, these genes play significant roles in immune regulation, cellular signaling, and protein homeostasis, with implications for immune responses and cancer biology.

Over half of HNSCC patients experience recurrence or metastasis despite current treatments, including immunotherapy, underscoring the need for new targeted therapies ^44^. The poor performance of *CD226*, *CD38*, and *KBTBD8* as markers in differentiating the three HNSCC clusters in the TCGA cohort highlights the need for new marker genes to improve classification. This discrepancy between OSCC-GB and HNSCC could be due to the unique profile of a subset of TCGA-HNSC patients known as HN2 subtypes. Markers specific to TCGA-HNSC have oncogenic roles in various cancers. For example, leupaxin (LPXN), involved in focal adhesion, promotes the proliferation, migration, and invasion of bladder cancer cells^45^. The transcription factor ETV7 was upregulated in breast and colorectal cancers ^46,47^. It stimulates the proliferation, migration, and cell cycle progression and suppresses the apoptosis in colorectal cancer cells ^47^.

Based on CD226, CD38, and KBTBD8 expression, GSE85195 patients were confidently classified within the GB1 patient cluster. Principal Component Analysis further confirmed that GSE85195 samples aligned closely with GB1 OSCC-GB patients. Additionally, these patients showed lower enrichment in immune-related gene sets and reduced levels of naive B cells, M1 macrophages, and CD4+ and CD8+ T-cells compared to the GB2 group. These findings underscore shared immunological and gene expression characteristics, suggesting potential avenues for targeted therapies in OSCC-GB patients.

In the TCGA-HNSC dataset, patients with predicted subtypes exhibit immune profiles similar to those observed in 41 patients with paired tumor-normal samples. HN3 tumors show the highest enrichment of immune cells, while HN2 tumors have the lowest enrichment. Patients in the HN1 and HN3 subtypes demonstrate better progress-free survival than those in the HN2 subtype. Moreover, HPV-positive patients may have a lower risk of cancer progression than HPV-negative TCGA-HNSC patients, possibly due to higher immune cell type scores in HPV-positive tumors ^48^. Like OSCC-GB patients, the highest levels of M1 macrophage and cytotoxic T-cell infiltrations in the tumor microenvironment were observed in HN3 subtypes of TCGA-HNSC patients, while the lowest levels were observed in HN2 tumors. HN1 and HN3 subtypes also exhibit significantly higher expression of MHC I molecules than HN2. Regardless of the anatomical location of tumors in the head and neck regions, PD-1/PD-L1(2), CTLA4/CD80(86), and BTLA/HVEM may be considered potential targets for treating immunologically hot sets of GB2 (or HN1 and HN3) tumors with checkpoint inhibition therapy.

In summary, gene set enrichment analysis on transcriptome data of OSCC-GB patients revealed activation of epithelial-mesenchymal transition, angiogenesis, and cell cycle functions, and significant involvement of the Warburg effect and altered fatty acid metabolism in tumors. Immune profiling showed significant enrichment of immune-related genes and immune cells, indicating the potential of immune checkpoint inhibition therapy. Comparison with TCGA-HNSC data revealed similar gene set enrichments and immune profiles. Some TCGA-HNSC patients showed subtle differences from OSCC-GB patients, indicating molecular heterogeneity of cancer of the head and neck region.

## Authors’ Contributions

Conceptualization: DD, PPM. Funding acquisition and project administration: RS, PPM. Data analysis: DD, PPM. Investigation: DD, PPM. Methodology: DD, AM, PPM. Sample acquisition: CKP, BR, RS. Sample examination and evaluation: SG. Data acquisition: AM. Writing of original draft: DD, PPM. Review and editing of draft: all authors.

### Funding

The Department of Biotechnology, Government of India, through the International Cancer Genome Consortium—India project and the Systems Medicine Cluster project.

## Acknowledgements

We acknowledge the financial support the Department of Biotechnology, Government of India, provides through grants under the International Cancer Genome Consortium— India project and the Systems Medicine Cluster project. PPM also extends his gratitude for the support from his National Science Chair fellowship, awarded by the Science & Engineering Research Board, Government of India.

## Data availability

The aligned BAM files for RNA-Seq data from the 72 tumor-normal paired samples can be accessed from the European Genome-phenome Archive (EGA) under study ID EGAS00001003893. The BAM files for RNA-Seq data from the 25 oral leukoplakia samples are also available from the European Nucleotide Archive (ENA) under study accession PRJEB42982.

